# Association Between moleculars and Osteoporotic Fracture Risk:A systematical review

**DOI:** 10.1101/2020.03.31.20049429

**Authors:** Jie-Yu Liu, Jia-Xiang Wang, Li Xu, Shu-Feng Lei, Fei-Yan Deng

## Abstract

Osteoporosis is a systemic chronic skeletal disease, which is characterized by low bone mineral density (BMD) and increased risk to osteoporotic fractures (OFs). OFs are associated with high mortality and morbidity, and seriously affect the life quality of patients. Osteoporosis is prevalent in the middle-aged and elderly population, especially the postmenopausal women. With population aging, osteoporosis becomes a world-wide serious public health problem. Early recognition of the high-risk population followed by timely and efficient intervention and/or treatment is important for preventing OFs. In light of the high heritability and complex pathogenesis of OP, comprehensive consideration of significant biological/biochemical factors is necessary for accurate risk evaluation. For this purpose, we reviewed recent research progress on moleculars which are diagnostic and/or predictive of OFs risk. Future integrative analyses and systematic evaluation of these moleculars may facilitate developing novel methodologies and/or test strategies, i.e., biochips, for early recognition of osteoporosis, hence to contribute to preventing OFs in the world.

**Graphical Abstract:** Osteoporosis, which is characterized by low bone mineral density (BMD) and increased risk to osteoporotic fractures (OFs), is prevalent in the middle-aged and elderly population, especially in the postmenopausal women. We focused on several types of important molecules, including proteins/peptides, RNAs, lipids, to gain comprehensive understanding and to generate novel perspectives in predicting and diagnosing OFs.

## Introduction

Osteoporosis is a chronic skeletal disease, which is characterized by low bone mineral density (BMD) and high risk to osteoporotic fracture (OF). Osteoporosis is prevalent in the middle-aged and elderly population, especially in the postmenopausal women. With population aging, osteoporosis and OFs have become an increasingly serious public health problem. A report from World Health Organization (WHO) in 2015 noted that OFs place a heavy burden on individuals and have important health consequences in both developed and developing countries [1].

A commonly used method to predict fracture risk is the FRAX tool [2], which has been modified or simplified in various countries and regions. Since the method was released, no molecular information has been incorporated into the prediction model thus far. A molecular, which was discovered by clinical test, is mainly composed of proteins, followed by small molecules and cells, and is measured as an indicator of normal biological or pathogenic processes, or a response to an exposure or intervention(FDA-NIH: molecular-Working-Group,2016).

The 10-year risks to hip fracture and major OF, which were calculated using the Chinese FRAX® model by the National Osteoporosis Foundation (NOF), suggested that the risk to OF increases with age in Chinese[3].A recent report showed that the FRAX method dramatically underestimate the fracture risk of Chinese population [4], which calls for more significant factors to be incorporated and more fine-tuned method to be applied for Chinese population for the purpose of precision prediction/medicine.

Osteoporosis results from imbalanced bone remodeling favoring bone loss due to relatively increased bone resorption by osteoclasts and/or decreased bone formation by osteoblasts. OFs are determined by both genetic and environmental factors, as well as their interactions. As reviewed previously, a large number of genes have been identified to be associated with BMD variation and OFs in humans [5, 6]. Meanwhile, rapid progress has been made in identification of novel biomolecules associated with OFs risks [7]. Therefore, it is reasonable for us to believe that targeted evaluation of a panel of significant biomolecules would become feasible in the near future, and facilitate developing effective moleculars to be applied for OF risk prediction.

For the above purpose and perspective, here we reviewed recent research progress on moleculars diagnostic and/or predictive of OFs, as follows.

## Methods

Since 1979, the number of publications on OF moleculars increases dramatically year by year, showing that continuous attention has been cast from investigators to the research field. To obtain a comprehensive collection of publications on moleculars of OFs, we performed an unrestricted PubMed search specifying co-occurrence of the terms “osteoporotic fractures and molecular”, “osteoporotic fractures and protein”, “osteoporotic fractures and RNA”, “osteoporotic fractures and metabolite”, “osteoporotic fractures and lipid”, then collected 15,781 publications.

To gain a comprehensive understanding and to generate novel perspectives, we reviewed the research progress in moleculars diagnostic and/or predictive of OFs. Considering that quite many reviews have already summarized the research progress at gene/SNP level, e.g., from genome-wide association and meta-analyses [5, 6], in this review, we focused on summarizing the progress on other types of molecules, such as proteins/peptides, RNAs, lipids, etc. As follows, the predictive and diagnostic moleculars were categorized and presented, respectively. However, this review based on the population of epidemical research, further studies and trials are needed to demonstrate the clinical values and the possible to be target of these novel moleculars.

### Potential predictive moleculars of OFs

Generally speaking, recent studies identified many circulating moleculars for OFs in the elderly. A majority are proteins which are predictive of OFs[8], such as insulin-like growth factor (IGF), periostin (POSTN), etc. In addition, more and more evidences suggest that metabolites, including nucleic acid, amino acid and lipid mediators, could predict OFs risk, as well. To date, further clinical application for these novel moleculars should be explained in clinical trials.

#### 1. Protein and peptide moleculars

A dozen of predictive protein moleculars associated with incident OF have been identified from prospective studies listed in **Table 1**. Risk predictive protein moleculars, including IGF binding protein-1(IGFBP-1)[9], periostin [10, 11], adiponectin[12, 13], C-reactive protein (CRP) [14], bone-specific alkaline phosphatase (BAP) [15] and fibroblast growth factor 23 (FGF23) [16, 17], were all positively associated with incident OFs.

**Table 1:**
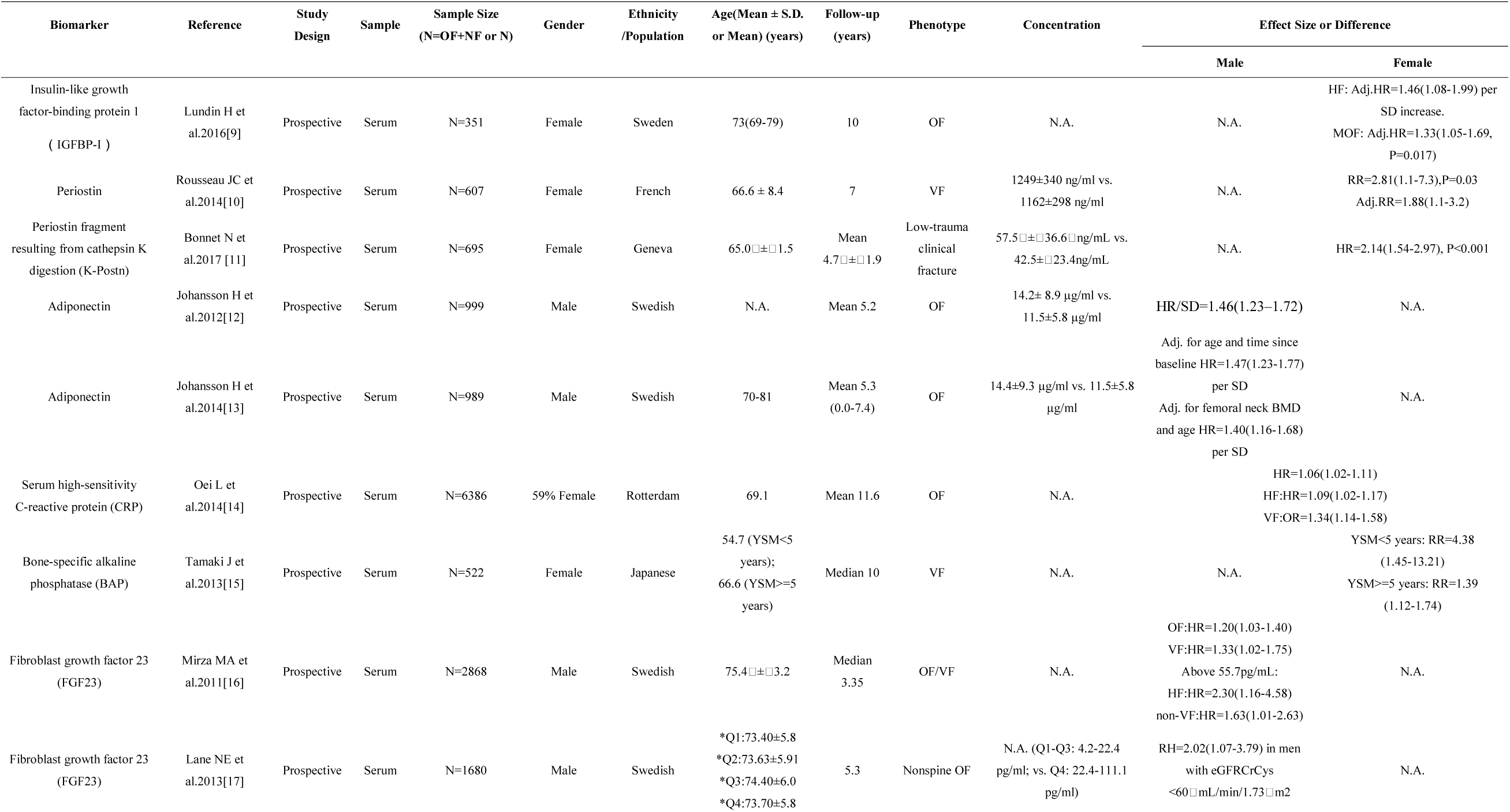
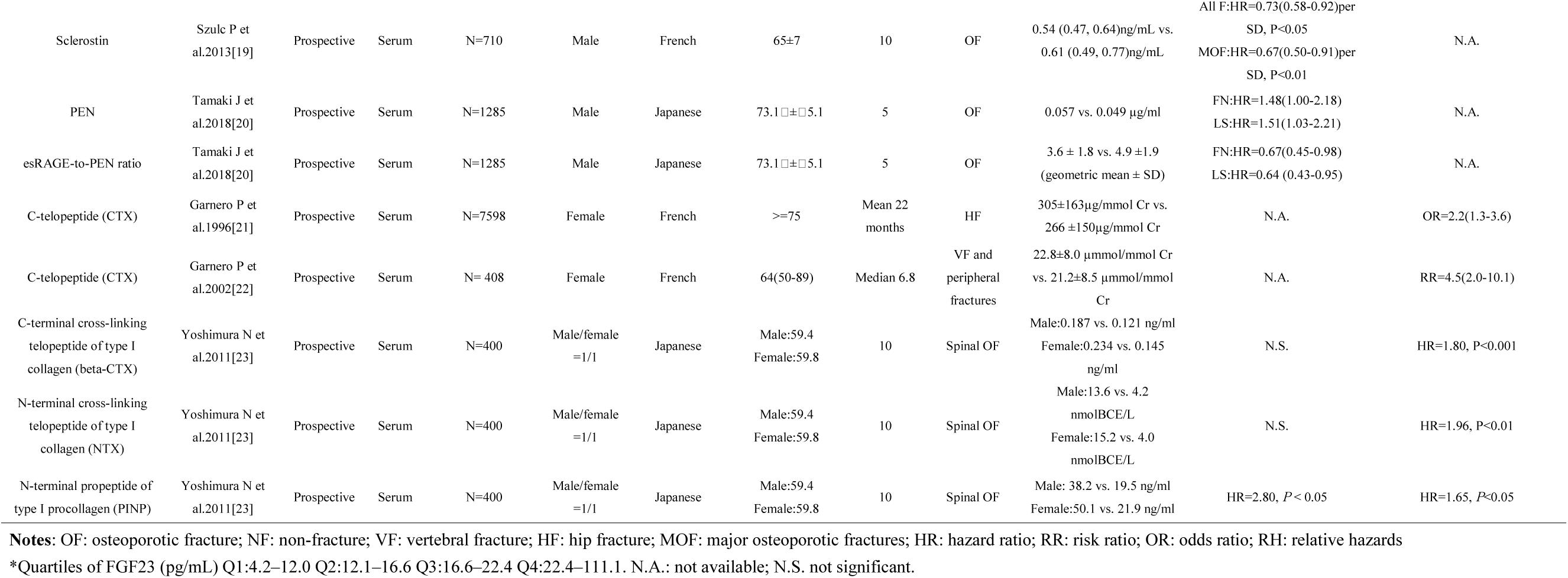
Predictive Protein and Peptide moleculars of Osteoporotic Fractures

Lundin H *et al*. discovered that IGF binding protein-1 (IGFBP-1) protein level in serum was positively correlated with the risk of hip fractures [Adj. hazard ratio(HR)=1.46 (1.08-1.99) per SD increase] and major OFs [HR=1.33 (1.05–1.69), *P*=0.017] in a population of 351 Sweden women [9]. Periostin is a secreted γ -carboxyglutamic acid-containing protein, which is expressed mainly in the periosteum of adult individuals[18]. Higher level of periostin in serum increases the fracture risk [10, 11]. For instance, Rousseau JC *et al*. discovered that the highest quartile of serum periostin was associated with an increased risk of incident fracture in a prospective study of 607 postmenopausal women [relative ratio (RR)=1.88 (1.1-3.2)] after adjustment for age, fragility fracture, and hip BMD T-score[10]. Periostin fragment resulting from cathepsin K digestion predicted OF risk in females as well [HR=2.14(1.54-2.97)] [11]. In 2012, Johansson H *et al*. found that the OF risk increased in parallel with increasing serum adiponectin [HR/SD=1.46(1.23–1.72)] and persisted after multivariate-adjusted analysis [HR/SD=1.30(1.09–1.55)] in 999 elderly Swedish males[12]. In 2014, Johansson H *et al*. highlighted the predictve value of serum adiponectin for OF in males [HR=1.47(1.23-1.77) per SD after adjusted for age and time since baseline, and HR=1.40(1.16-1.68) per SD after adjusted for femoral neck BMD and age][13].

Besides, the serum CRP level was associated with a risk to any type of OFs [HR=1.06(1.02-1.11)], hip fractures (HF) [HR=1.09(1.02-1.17)] and vertebral fractures (VF)[OR=1.34(1.14-1.58)] in a prospective study of 6,386 participants (59% female) [14]. However, the clinical value of CRP and its potential diagnostic ability for OF need to be further validated, especially in single patient, since we only explained its associations based on epidemical research. Serum BAP was also reported to predict an increased risk of osteoporotic VF [RR per SD=4.38(1.45–13.21)] in 65 postmenopausal women with years of menopause (YSM) <5 years. In 457 postmenopausal women with YSM ≥ 5 years, the upper tertile for BAP had a greater risk of vertebral fractures[RR=1.39(1.12-1.74)][15]. Furthermore, Mirza MA *et al*. pointed that FGF23 protein level in serum above 55.7□ pg/mL was associated with an increased risk to HF [HR=□ 2.30(1.16-4.58)] and non-VF [HR=□ 1.63(1.01-2.63)], respectively in a population-based OF study in men (MrOS; N=2868), and subjects in the highest quartile of serum FGF23 protein level (>57.4 pg/ml) were at a 63% increased risk to non-vertebral OFs, compared to those in the lowest three quartiles [16]. Lane NE *et al*. discovered that in men with poor renal function only (eGFR_CrCys_< 60ml/min/1.73m^2^), the relative hazards (RH) of hip and non-spine OFs was 2.02 (95% CI=1.07-3.79) in the highest quartile of serum FGF23 protein level, as compared to the rest [17].

Besides, a couple of protective protein moleculars for OFs, including sclerostin [19] and the endogenous secretory receptor for Advanced Glycation End Products (esRAGE)-to-Pentosidine (PEN) ratio [20], have been identified thus far. For example, Szulc P*et al*. reported that French men with higher concentration of serum sclerostin had lower OFs risk [HR□ =□ 0.55(0.31-0.96), *P*□ <□ 0.05 adjusted for hip BMD][19]. In addition, theesRAGE was associated with reduced activity of PEN [20]. Tamaki J *et al*. found that the esRAGE-to-PEN ratio was negatively correlated with incident OFs in 1,285 elderly male Japanese [(HR (95%Cl)= 0.67 (0.45-0.98) and 0.64 (0.43-0.95), adjusted for age, glycated hemoglobin A1c, and T-score of BMD at FN or LS] [20].

As for the peptide moleculars, such as bone resorption markers CTX [21-23] and bone formation marker PINP [23, 24], showed great predictive values for OFs. For example, Early in 1996, Garnero P *et al*. found that CTX level above the upper limit of the premenopausal range was associated with an increased risk of hip OF in a population of 7,598 healthy French [OR=2.2(1.3-3.6)][21]. In 2002, Garnero P *et al*. found that women with both increased urinaryalpha-L/beta-L-CTX ratio and low femoral neck BMD (T score<-2.5) had a higher risk of OFs[RR=4.5(2.0–10.1)] in a prospective study [22]. Yoshimura N *et al*. reported that serum beta-CTX and NTX was significantly related to the occurrence of spinal OFs in Japanese women (beta-CTX: HR=1.80, *P*<0.001; NTX: HR=1.96, *P*<0.01) after adjusting for confounders in multivariate analysis [23], and serum PINP level was significantly higher in OFs than controls in both men (HR=2.80, *P*<0.05) and women (HR=1.65, *P*<0.05) [23].

#### 2. Potential Metabolite moleculars

Metabolites play an essential role in predicting OFs. Some polar metabolites are significantly involved in nucleic acid metabolism, amino acid metabolism and lipid metabolism, which are closely related to the imbalance between bone resorption and formation and may underlie the development of OFs [25]. Predictive metabolite moleculars associated with OFs risk were summarized in **Table 2** and presented in the following three categories.

**Table 2:** Predictive Metabolite moleculars of Osteoporotic Fractures

| Biomarker | Reference | Study Design | Sample | Sample Size<br>(N=OF+NF or N) | Gender | Ethnicity<br>/Population | Age(Mean ± S.D. or Mean)<br>(years) | Follow-up<br>(years) | Phenotype | Concentration | Effect Size or Difference |  |
| --- | --- | --- | --- | --- | --- | --- | --- | --- | --- | --- | --- | --- |
|  |  |  |  |  |  |  |  |  |  |  | Male | Female |
| free deoxypyridinoline | Garnero P et al.1996[21] | Prospective | Urine | N=7598 | Female | French | >=75 | Mean 22 months | OF | 6.41±2.20nmol/mmol<br>Cr vs.<br>5.80±1.94nmol/mmol<br>Cr | N.A. | OR=1.9(1.1-3.2) |
| urinary deoxypyridinoline | Yoshimura N et al.2011[23] | Prospective | Urine | N=400 | Male/female =1/1 | Japanese | Male:59.4<br>Female:59.8 | 10 | Spinal OF | Male:3.06 vs. 1.53<br>pmol/lmolCr<br>Female:4.76 vs. 1.91<br>pmol/lmolCr | N.S. | HR=1.40, P<0.05 |
| homocysteine | Gjesdal CG et al.2007[26] | Prospective | Plasma | N=4766 | Male/female =2127/2639 | Bergen | 65-67 years at enrollment | Median 12.6 | HF | 7–20 μM | N.S. | HR=2.42(1.43-4.09) |
| homocysteine | Enneman AW et al.2012[27] | Prospective | Plasma | N=503 | Female | Rotterdam | 68.5(61.3–74.9) | Mean 7 | OF | (9.4–11.6)μmol/L | N.A. | Adj. for age, BMI, FN-BMD.<br>HR=1.82(1.02-3.22) |
| Dimethylglycine(DMG) | Øyen J et al.2015[28] | Longitudinal | Plasma | N=3310 | Male/female =1017/1204 | Bergen, Norway | 71-74 | Mean 10.8 | HF | (4.4±1.5)μmol/L | Adj. for BMD, HR=1.70(1.28-2.26) |  |
| Sphingosine-1-phosphate (S1P) | Ardawi MM et al.2018[32] | Prospective longitudinal | Plasma | N=707 | Female | Saudi Arabia | 61(53-93) | Mean (5.2±1.3) | OF | 7.23±0.79 μmol/L<br>vs.5.02±0.51 μmol/L | N.A. | Adj.HR=6.12(4.92-7.66) per SD increase. |
| 24,25-dihydroxyvitmin D [24,25(OH)2D] | Ginsberg C et al.2018[33] | Longitudinal | Serum | N=890 | Male/female =267/178 | 16%<br>African-American | 78 | Mean 8.4 | HF | (1.7±1.0) ng/ml | HR=0.73(0.61-0.87) per SD increase |  |
| the ratio of 24,25(OH)2D to 25(OH)D [VMR] | Ginsberg C et al.2018[33] | Longitudinal | Serum | N=890 | Male/female = 267/178 | 16%<br>African-American | 78 | Mean 8.4 | HF | (6.84±2.23) ng/ml | HR=0.74(0.61-0.88) per SD increase |  |
| 25-hydroxyvitaminD(25OHD) | Swanson CM et al.2015[34] | Prospective | Serum | N=1000 | Male | USA | 74.6±6.2 | Mean 5.1 | non-VF | 3.13 vs. 20.90<br>ng/mL | HR=2.01(1.24-3.26) | N.A. |
| 1,25-dihydroxyvitaminD (1,25(OH)2D) | Swanson CM et al.2015[34] | Prospective | Serum | N=1000 | Male | USA | 74.6±6.2 | Mean 5.1 | non-VF | 8.70 vs. 51.60 pg/mL | HR=1.99(1.19-3.33) | N.A. |
| Polyunsaturated fatty acids (PUFAs) | Harris TB et al.2015[37] | Longitudinal | Plasma | N=540+898 | OF:male/fe<br>male =<br>19/35<br>NF:<br>male/female<br>=429/469 | N.A. | OF:78.5± 5.69<br>NF:76.5± 5.48 | Mean 7 | OF | N.A. | HR=0.60(0.41-0.89) | Inverse association, P-trend = 0.06 |

Notes: OF: osteoporotic fracture; NF: non-fracture; VF: vertebral fracture; HF: hip fracture; FN: femoral neck; HR: hazard ratio; OR: odds ratio.
\*N.A.: not available; N.S. not significant.

##### 2.1 Nucleic acid metabolite

Some nucleic acid metabolite, play a key role in the bone metabolism. According to the EPIDOS cohort study including 7,598 female French, high level of urine free deoxypyridinoline was associated with increased risk of HFs, and free deoxypyridinoline secretion above the upper limit of the premenopausal range was also associated with an increased HFs risk [OR=1.9(1.1-3.2)][21]. Yoshimura N *et al*. found that high level of urinary deoxypyridinoline significantly predicted 10-year OFs risk at the lumbar spine in women (HR=1.40, P<0.05) but not in men [HR=1.53(0.63-3.73)][23]. To be honest, the significant associations were only validated in epidemical research, rather than in specific clinical treatment, especially for single patient, so the potiential predictive value of them needs to be addressed.

##### 2.2 Amino acid moleculars

Plasma total homocysteine(tHcy) was frequently reported to be associated with OFs risk [26, 27]. The adjusted HR (95% CI) for OFs in subjects with high (>=15 microM) vs. low levels (<9.0 microM) of tHcy was significant in women [HR=2.42 (1.43-4.09)] but not in men [HR=1.37 (0.63-2.98)] ina prospective study of 4,776 elderly [26]. Whereas incidence of OFs was associated with quartiles of homocysteine (Hcy) (*P*=0.047), higher risks of OFs was observed in females aged 55 years and over [adjusted for age, BMI, FN-BMD, HR=1.82(1.02-3.22)] from the Rotterdam Study (RS) in a prospective study of 7 following-up years[27].

Dimethylglycine (DMG) is a product of the choline oxidation pathway and generated from betaine during the folate-independent remethylation of Hcy to methionine. Lower level of plasma DMG was associated with an increased risk of incident HF [HR=1.70(1.28-2.26)], as compared to the highest tertile[28].

##### 2.3 Lipid mediators

Osteoporosis or osteopenia often co-exist with lipid metabolism disorders [29]. Studies suggested that peroxisome proliferator activated receptor gamma (PPARγ) may play a key role in the pathogenesis of both osteoporosis and lipid metabolism disorder, and lipid metabolism disorders caused by bone marrow fat may be the main cause of bone marrow microcirculation disorders and osteopenia [30].

Sphingosine-1-phosphate (S1P) is a lipid mediator which acts through S1P receptors.S1Pplays diverse roles in cell differentiation, apoptosis, proliferation, motility, bone metabolism, and the most prominent role seems to be the augmentation of bone resorption [31]. In another prospective longitudinal study of 707 postmenopausal women, the plasma S1P levels were significantly higher in the women with incident OFs than in those without osteoporosis related fractures (ORFs) (7.23±0.79 µmol/L vs. 5.02±0.51 µmol/L; *P*<0.001) [32].After adjustment for age and other confounders, the HR of OFs was 6.12 (95% CI: 4.92-7.66) per SD increase in plasma S1P level and the women in the highest quartile of S1P levels had a significantly increased fracture risk [HR=9.89(2.83-34.44)][32].

Ginsberg C *et al*. revealed the association of vitamin D with hip BMD and the risk of HF among 890 participants followed-up for ∼8.4-years, both higher 24,25(OH)_2_D and vitamin D metabolite ratio (VMR, i.e., the ratio of 24,25(OH)_2_D to 25-hydroxyvitaminD (25OHD)) were associated with lower risk of HF [HR=0.73(0.61-0.87)] and 0.74 (0.61-0.88) per SD increase, respectively] [33]. Swanson CM *et al*. also reported that 25OHD and 1,25(OH)_2_D were protective against HFs [34]. However, previous studies suggested that the hip fracture protection found with higher 25OHD levels is largely mediated by BMD[35]. Specifically, the risk of HFs was higher in men with the lowest 1,25(OH)_2_D levels (8.70 to 51.60 pg/mL) after adjustment for baseline hip BMD [HR (95%Cl)=1.99(1.19–3.33)]; and significantly increased risk of HFs was also observed for the lowest quartile of 25OHD (3.13 to 20.90 ng/mL) in American man [HR=2.01(1.24-3.26)][34]. However, there existed some inconsistent results regarding to the associations between 25(OH)D and high risk of HF[36]. To our knowledge, no prior study has evaluated whether 24,25(OH)_2_D concentrations or the VMR are more strongly associated with bone health than 25(OH)D alone. Additionally, the associations of 25OHD and 1,25 (OH)2D with baseline BMD and BMD change were independent of each other. These results do not support the hypothesis that measures of 1,25(OH)2D improve the ability to predict adverse skeletal outcomes when 25OHD measures are available. Still, there is long way before reaching the potential diagnostic role of 25OHD, 1,25(OH)_2_D and risks to OFs. Epidemical outcomes need to be further elaborated when apply to clinical trials.

Besides, Harris TB *et al*. reported that higher concentration of plasma polyunsaturated fatty acids (PUFAs) was associated with lower OFs risk in older adults [37]. Among 1,438 participants, 540 participants with incident OFs, the highest tertile of plasma PUFAs was associated with decreased OFs risks [HR=0.60(0.41-0.89)] in men; PUFAs tended to be inversely associated with OFs risks in women (*P*_*trend*_ = 0.06)[37]. However, PUFAs are novel moleculars related to OFs and only conducted in epidemcal research among population, the further significant value in clinical usage to treat OFs need to be validated in single patient.

### Potential Diagnostic moleculars of OFs

Some moleculars, such as insulin-like growth factor-1 (IGF-1), and IGF-binding protein-3 (IGFBP-3), etc. demonstrated great diagnostic values for OF. Recently, more scientific research suggested that microRNAs (miRNAs) and metabolites are diagnostic of OFs risk, as well.

#### 1. Potential protein moleculars

Potential diagnostic protein moleculars associated with OF risks were identified in cross-sectional studies, which were summarized and listed in **Table 3**. In 1997, Sugimoto T *et al*. reported that serum IGF-I protein level was positively correlated with BMD and negatively associated with spinal OF in Japanese women [38]. Nakaoka D *et al*. also discovered that IGF-I protein was negatively correlated with the risk of spinal OFs in Japanese postmenopausal women [HR=0.19(0.07-0.59)][39]. Similar results were reported by Yamaguchi T *et al*. for VF [OR=0.29 (0.15-0.57) per SD increase, *P*=0.0003] in Japanese postmenopausal women [40]. Besides, IGF-binding protein-3 (IGFBP-3) was also negatively associated with both spinal OFs (r=-0.409, *P*<0.0001) [38] and VF [OR=0.31(0.16-0.61), *P*=0.0007][40] in Japanese women. As reported by Kim BJ *et al*., plasma periostin was a risk factor of any fracture [OR=1.50(1.14-1.97), *P*=0.003], non-VF fracture [OR=1.59(1.12-2.24), *P*=0.009]in Koreans [41]. The odds for non-VF was 2.48-fold higher in the highest vs. lowest tertiles of plasma periostin level (95%Cl=1.10-5.61)[41]. Consistently, Yan *et al*. reported that serum periostin level was positively correlated with femoral neck BMD (r=-0.529, *P*<0.001) in Chinese elderly females[42], supporting that it is a risk marker of OFs.

**Table 3:** Potential Diagnostic Protein moleculars of Osteoporotic Fractures

| Biomarker | Reference | Study Design | Sample | Sample Size<br>(N=OF+NF or N) | Gender | Ethnicity/<br>Population | Age(Mean ± S.D.<br>or Mean) (years) | Phenotype | Concentration | Effect Size or Difference |
| --- | --- | --- | --- | --- | --- | --- | --- | --- | --- | --- |
| IGF-I | Sugimoto T et al.1997[38] | Cross-sectional | Serum | N=165 | Female | Japanese | 62 | Spinal OF | N.A. | Lower in OF, P<0.01; Cutoff 110 ng/ml provided a specificity of 81% with a sensitivity of 76%. |
| IGF-I | Nakaoka D et al.2001[39] | Cross-sectional | Serum | N=205 | Female | Japanese | 64 | Spinal OF | N.A. | OR=0.19(0.07-0.59), P=0.003 |
| IGF-I | Yamaguchi T et al.2006[40] | Cross-sectional | Serum | N=61+132 | Female | Japanese | 62.5(46-88) | VF | 97.1±32.1 ng/dL vs. 143.9±40.9 ng/dL | OR=0.29(0.15-0.57), P=0.0003 |
| IGFBP-3 | Sugimoto T et al.1997[38] | Cross-sectional | Serum | N=165 | Female | Japanese | 62 | Spinal OF | N.A. | Lower in OF, P<0.01; Cutoff 2.1µg/ml provided a specificity of 81% with a sensitivity of 81%. |
| IGFBP-3 | Yamaguchi T et al.2006[40] | Cross-sectional | Serum | N==61+132 | Female | Japanese | 62.5 (46-88) | VF | 2.18±1.02 µg/mL vs. 3.23±1.07 µg/mL | OR=0.31(0.16-0.61), P=0.0007 |
| Periostin | Kim BJ et al.2015[41] | Case-control | Plasma | N==133+133 | Female | Korean | 62.7±6.4 Cases;<br>63.0±6.3 Control | OF | 32.4 ng/mL vs. 29.2 ng/mL, any fracture;<br>32.6 ng/mL vs. 28.6 ng/mL, non-VF. | Any fracture: OR=1.50(1.14-1.97), P=0.003<br>Non-VF: OR=1.59(1.12-2.24), P=0.009 |
| Periostin | Yan J et al.2017[42] | Case-control | Serum | N==261+106 | Female | Chinese | 80 (76–84) Cases;<br>78 (72–83) Controls | HF | 45.51 (28.77–79.70) ng/mL vs. 32.87 (19.71–53.58) ng/mL | FN-BMD: β(standardized coefficient)=−0.390,P=0.004<br>LS-BMD: β(standardized coefficient)=−0.158,P=0.190 |

**Notes:** OF: osteoporotic fracture; NF: non-fracture; VF: vertebral fracture; HF: hip fracture; LS: lumbar spine; FN: femoral neck; OR: odds ratio.
\*N.A.: not available; N.S. not significant.

#### 2. Potential MicroRNA moleculars

MicroRNA (miRNAs) is a class of small (∼22 nucleotides), single-stranded noncoding RNAs. They are considered “micro-managers” of gene expression, and their role in regulating biological functions has been increasingly recognized. The diagnostic miRNAs moleculars associated with OFs risk were summarized in **Table 4**.

**Table 4:** Diagnostic microRNAs moleculars of Osteoporotic Fractures

| microRNA ID | Reference | Study Design | Sample Size (N=OF+NF or N) | Sample | Expression Trend in Case vs. Control | Fracture Diagnostic Value or Difference | Phenotype |
| --- | --- | --- | --- | --- | --- | --- | --- |
| miR-21 | Claudine Seeliger et al.2014[43] | Case-control | Serum:N=30+30<br>Bone:N=20+20 | Serum and Bone tissue | Up-regulated | AUC:0.63(0.53–0.73;P= 0.013) | OF |
| miR-23a | Claudine Seeliger et al.2014[43] | Case-control | Serum:N=30+30<br>Bone:N=20+20 | Serum and Bone tissue | Up-regulated | AUC:0.63( 0.53–0.73;P=0.015) | OF |
| miR-24 | Claudine Seeliger et al.2014[43] | Case-control | Serum:N=30+30<br>Bone:N=20+20 | Serum and Bone tissue | Up-regulated | AUC:0.63 (0.53–0.74;P=0.013) | OF |
| miR-93 | Claudine Seeliger et al.2014[43] | Case-control | Serum:N=30+30<br>Bone:N=20+20 | Serum | Up-regulated | AUC:0.68 (0.58–0.78;P=0.001) | OF |
| miR-100 | Claudine Seeliger et al.2014[43] | Case-control | Serum:N=30+30<br>Bone:N=20+20 | Serum and Bone tissue | Up-regulated | AUC:0.69(0.60–0.78;P=0.0003) | OF |
| miR122a | Claudine Seeliger et al.2014[43] | Case-control | Serum:N=30+30<br>Bone:N=20+20 | Serum | Up-regulated | AUC:0.77(0.69–0.86,P<0.0001) | OF |
| miR-124a | Claudine Seeliger et al.2014[43] | Case-control | Serum:N=30+30<br>Bone:N=20+20 | Serum | Up-regulated | AUC:0.69(0.59–0.78;P=0.0005) | OF |
| miR-125b | Claudine Seeliger et al.2014[43] | Case-control | Serum:N=30+30<br>Bone:N=20+20 | Serum | Up-regulated | AUC:0.76( 0.67–0.85;P< 0.0001) | OF |
| miR-148a | Claudine Seeliger et al.2014[43] | Case-control | Serum:N=30+30<br>Bone:N=20+20 | Serum | Up-regulated | AUC:0.61 (0.51–0.72;P=0.038) | OF |
| miR-21-5p | Layla Panach et al.2015[44] | Case-control | N=15+12 | Serum | Up-regulated | Fold change=1.57, P=0.002 | OF |
| miR-122-5p | Layla Panach et al.2015[44] | Case-control | N=15+12 | Serum | Up-regulated | Fold change=5.48, P=0.00008 | OF |
| miR-125b-5p | Layla Panach et al.2015[44] | Case-control | N=15+12 | Serum | Up-regulated | Fold change=4.62, P=0.0003 | OF |
| miR-328-3p | Sylvia Weilner et al.2015[46] | Cross-sectional | N=37 | Serum | Down-regulated | Fold change=-1.54, -2.04; P<0.05 | OF |
| let-7g-5p | Sylvia Weilner et al.2015[46] | Cross-sectional | N=37 | Serum | Down-regulated | Fold change=-1.24, -1.89; P<0.05 | OF |
| miR-19a-3p | Kocijan R et al.2016[47] | Case-control | N=36+39 | Serum | Down-regulated | AUC:0.929 (0.856–0.983) | Low traumatic fractures |
| miR-19b-3p | Kocijan R et al.2016[47] | Case-control | N=36+39 | Serum | Down-regulated | AUC:0.944 (0.879–0.997) | Low traumatic fractures |
| miR-30e-5p | Kocijan R et al.2016[47] | Case-control | N=36+39 | Serum | Down-regulated | AUC:0.959 (0.901–0.997) | Low traumatic fractures |
| miR-140-5p | Kocijan R et al.2016[47] | Case-control | N=36+39 | Serum | Down-regulated | AUC:0.947 (0.900–0.983) | Low traumatic fractures |
| miR-152-3p | Kocijan R et al.2016[47] | Case-control | N=36+39 | Serum | Down-regulated | AUC:0.962 (0.918–0.993) | Low traumatic fractures |
| miR-324-3p | Kocijan R et al.2016[47] | Case-control | N=36+39 | Serum | Down-regulated | AUC:0.950 (0.885–0.994) | Low traumatic fractures |
| miR-335-5p | Kocijan R et al.2016[47] | Case-control | N=36+39 | Serum | Down-regulated | AUC:0.939 (0.872–0.986) | Low traumatic fractures |
| miR-550a-3p | Kocijan R et al.2016[47] | Case-control | N=36+39 | Serum | Down-regulated | AUC:0.909 (0.837–0.970) | Low traumatic fractures |
| hsa-miR-4516 | Mandourah AY et al.2018[50] | Case-control | N=139 | Serum and Plasma | Up-regulated | P = 0.00014 | OF |
| miR-140-3p | Ramírez-Salazar EG et al.2018[51] | Case-control | N=20+20 | Serum | Up-regulated | AUC:0.92;P< 0.0001 | OF |
| miR-23b-3p | Ramírez-Salazar EG et al.2018[51] | Case-control | N=20+20 | Serum | Up-regulated | AUC:0.88;P< 0.0001 | OF |

**Notes:** OF: osteoporotic fracture; NF: non-fracture; AUC: area under curve.

Early in 2014, Seeliger C *et al*. identified 9 serum miRNAs (miR-21, miR-23a, miR-24, miR-93, miR-100, miR-122a, miR-124a, miR-125b, and miR-148a) significantly up-regulated in OFs in a case-control study, which presented modest diagnostic performance individually (AUC=0.61-0.77, *P*<0.05). Among the above nine miRNAs, miR-21, miR-23a, miR-24, miR-100 displayed a significantly higher expression level in bone tissues in OFs compared with controls [43]. In 2015, Panach L*et al*. showed that 3 miRNAs in serum (miR-122-5p, miR-125b-5p and miR-21-5p) were up-regulated in fracture cases vs. controls [44]. Indeed, miR-21-5p was one of the few miRNAs, together with miR-223 and miR-155, which are directly related to osteoclast differentiation [45]. In an explorative analysis of 175 serum miRNAs in patients with recent OFs and age-matched control, the down-regulation of miR-328-3p and let-7g-5p in response to OF were identified (Fold Change (FC)=-1.54, *P*=0.034 and FC=-1.24, *P*=0.041, respectively) and later confirmed (FC=-2.04, *P*=0.016 and FC=-1.89, *P*=0.027, respectively) in an independent case-control sample[46].

Furthermore in 2016, Kocijan R *et al*. designed a case-control study including a cross-sectional design of 36 patients with prevalent low-traumatic fractures and 39 control subjects, one hundred eighty-seven miRNAs were quantified in serum by qPCR, compared between groups and correlated with established bone turnover markers. Eight serum miRNAs (miR-152-3p, miR-30e-5p, miR-140-5p, miR-324-3p, miR-19b-3p, miR-335-5p, miR-19a-3p, miR-550a-3p) were found down-regulated with low traumatic fractures, which excellently discriminated OF cases and controls, regardless of age and sex (AUC>0.9, *P*<0.05) [47]. In this way, miRNAs might be directly linked to bone tissue homeostasis because of the significant correlations between miR-29b-3p and P1NP[48], miR-365-5p and iPTH, TRAP5b, P1NP and Osteocalcin[49].

Mandourah AY*et al*. reported that the circulating levels of miR-4516 was significantly lower (*P*<0.001) in the OFs with low BMD than non-fracture subjects with low BMD or non-osteoporotic controls [50]. Significantly, Ramírez-Salazar EG *et al*. reported that serum miR-140-3p and miR-23b-3p, which were significantly elevated in OF vs. NF subjects, presented high performances in relation to OFs (AUC=0.92, *P*<0.0001; AUC=0.88, *P*<0.0001)[51].

To conclude, each miRNA has specific characteristics predicting risk of osteoporosis. For example, miR-214 inhibits bone formation via ATF4 and is down-regulated in osteoporotic bone. As concluded by

Chen et al. [52] there are biological differential expressions in some c-miRNAs between osteoporotic and non-osteoporotic individuals, but their study did not determine that these specific circulating miRNAs were moleculars of osteoporosis.

### 3. Potential metabolite moleculars

Metabolites, such as urinary mineral products, amino acid and lipid mediators, all play essential roles in bone metabolism as described above. Potential diagnostic metabolite moleculars of OFs were summarized and listed in **Table 5** and presented in the following three categories.

**Table 5:**
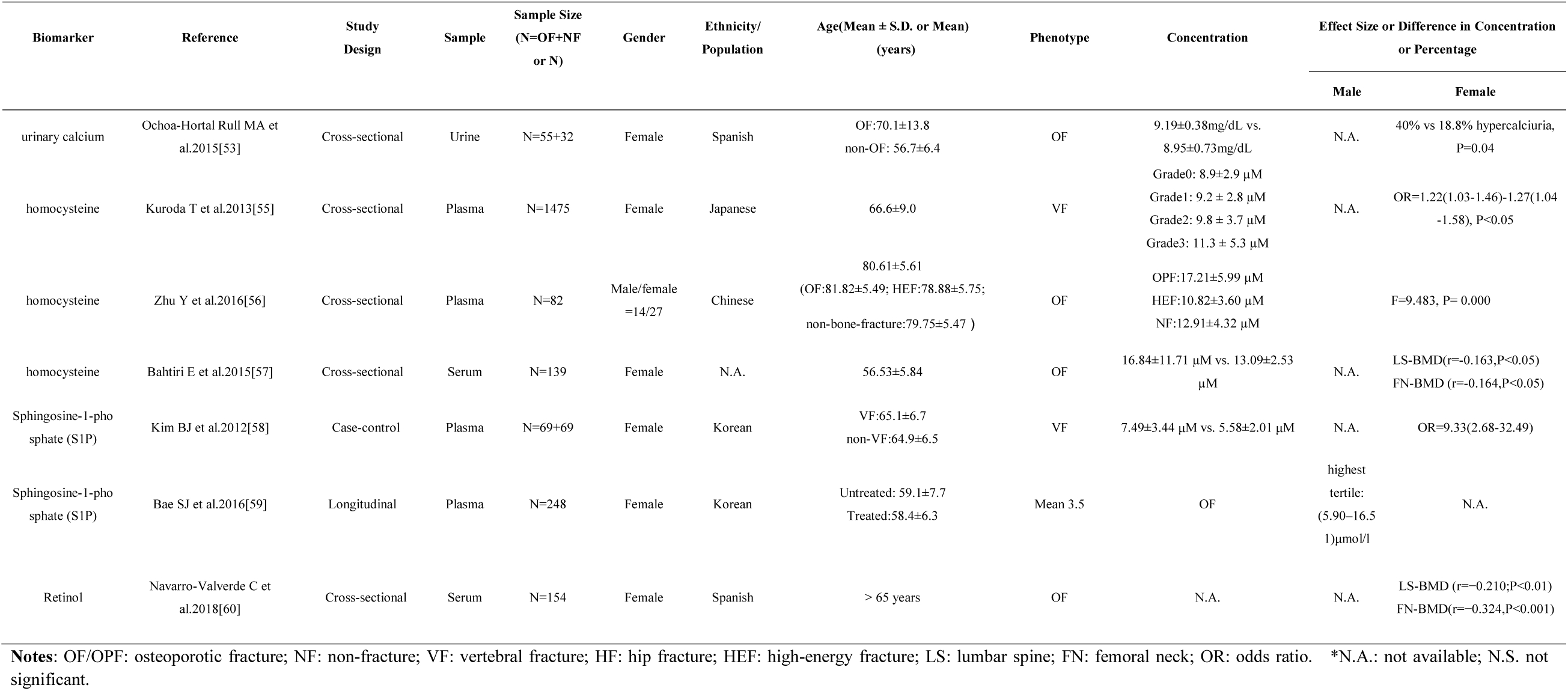
Potential Diagnostic Metabolite moleculars of Osteoporotic Fractures

#### 3.1 Urinary mineral moleculars

Elevated fasting urinary calcium/creatinine ratio mainly manifested the process of osteoporosis. The percentage of hypercalciuria in urine in OF women (40%) was significantly higher compared to non-OF women (18.8%) (*P*=0.04) in a Spanish cross-sectional study, as reported by Ochoa-Hortal Rull MA[53]. 24-hour urinary calcium and fasting calcium/creatinine ratio may show some great characters as potential diagnostic moleculars for OFs, to some extent [54]. However, these results were only obtained from epidemical studies, more researches demonstrated these values of clinical application are warrented.

#### 3.2 Amino acid moleculars

Homocysteine (Hcy) level in serum or plasma was a potential diagnostic molecular of OFs. Kuroda T *et al*. discovered that the plasma tHcy level was a significant risk factor for severe VFs in postmenopausal Japanese women [OR=1.22(1.03-1.46)-1.27(1.04-1.58), *P*<0.05) [55], and the plasma tHcy level was significantly higher in moderate VFs (Grade 1, 9.2 ± 2.8 µM) and severe VFs (Grade 2, 9.8 ± 3.7 µM), as compared to the Grade 0 (8.9±2.9 µM) [55].Besides, plasma Hcy concentration differed significantly between OF group (17.21±5.99 µM), high-energy fracture (HEF) group (10.82±3.60µM) and non-bone-fracture (NF) group (12.91±4.32µM) (F=9.483), and was positively correlated with bone resorption markers (serum C-terminal cross linking telopeptide of type I collagen (S-CTX), plasma levels of 25-OH Vit D) related to OF fracture in elderly Chinese cohort[56]. Consistently, Bahtiri E *et al*. reported that serum Hcy level was inversely associated with both lumbar spine BMD (r=-0.163, *P*<0.05) and femur neck BMD (r=-0.164, *P*<0.05), supporting it as a risk factor of OFs [57]. Of note, reducing plasma Hcy level can improve bone quality and reduce the risk of falling in elderly patients secondary to bone or skeletal muscle weakness, thereby reducing the risk of OPFs in the elderly. But how to predict the occurrence of OPFs is the direction for future research.

#### 3.3 Lipid mediators

Lipid mediators, such as S1P and retinol, also play an important role in diagnosing OFs. For instance, plasma S1P levels were markedly higher in subjects with VFs (7.49±3.44μM) than in those without VFs (5.58±2.01μM) [OR=9.33(2.68-32.49), *P*=0.001], and increased with the number of VFs in a dose-response manner (*P*_*for the trend*_ <0.001) [58]. An independent study reported that the adjusted OR (95%Cl) of plasma S1P levels for OFs was 2.05(1.03-4.00)[59]. Navarro-Valverde C*et al*. reported that the serum retinol level was a risk factor of OFs, which was negatively correlated with BMD at lumbar spine (r=-0.210; *P*<0.01) and femoral neck (r=-0.324, *P*<0.001) after multivariable adjustment [60]. A particularly interesting finding is that the plasma S1P level was associated with the risk of incident fracture, even after additional adjustment for antiosteoporotic medication. These results could be partially explained by the potential effects of S1P on the response to the antiosteoporotic medication. In conclusion, plasma S1P level, in combination with the BMD, could be a useful marker for predicting the risk of osteoporotic fracture.

## Conclusions and Future Prospective

OFs are associated with high mortality and morbidity, and seriously affect the life quality of patients worldwide. In this study, we systematically reviewed the international research progress on the other types of predictive and diagnostic moleculars for OFs, and summarized significant moleculars with potential clinical utility. Collectively, remarkable variations in protein, miRNA and metabolite levels with the risk of OFs were principally identified through case-control studies and/or longitudinal cohort studies in various ethnicities.

The achievements in OF molecular development, as reviewed above, from clinical observations or discoveries to the recent fundamental researches of bone biology, represent private and public attention, investments, and efforts towards understanding the pathogenesis and preventing the occurrence of OFs. As reported in many clinical trials, some protein moleculars, including IGFBP-1, periostin, adiponectin, CRP, BAP and FGF23, etc. are positively associated with incident OFs. Other proteins, such as IGF-1, IGFBP-3 and periostin, demonstrate great diagnostic values for OFs. Besides these significant protein moleculars, metabolite products, such as urinary mineral metabolite, amino acid and lipid mediators might be crucial markers, as well. Furthermore, we have also summarized some reliable miRNAs markers, which demonstrate great diagnostic values (AUC>0.9, *P*<0.05) in OFs.

In conclusion, great progress has been made in OF moleculars discovery in the past decade, and some novel and promising moleculars have emerged. To be noted, clinical data on these novel moleculars remain limited, and sometimes the epidemical results were controversial among independent study samples. For example,the miR-22-3p was found to be moderately up-regulated in the discovery cohort, but significant down-regulation was observed in the validation cohort[46].

Therefore, it is necessary and would be important to confirm the potential clinical utility of the novel moleculars through independent studies and with larger sample sizes. With global aging population increasing, senile osteoporosis and the resultant fractures incur heavy social and economic burden. The new discoveries with no doubt will greatly promote the development of reliable moleculars for OFs and inform development of novel tools and strategies for diagnosing, predicting, and preventing OP and OFs.

## Data Availability

All data is available online.

## Conflict of interest statement

No conflict of interest were reported by all authors.

### Acknowledgement

Jieyu Liu and Li Xu analysed and interpreted the data; Jiaxiang Wang was in the writing of the manuscript; Shufeng Lei and Feiyan Deng were in the decision to submit the manuscript for publication.

